# Prevalence of Non-obese Type 2 Diabetes in economically disadvantaged Indian rural populations

**DOI:** 10.1101/2020.09.21.20198598

**Authors:** Saptarshi Bej, Jit Sarkar, Saikat Biswas, Pabitra Mitra, Partha Chakrabarti, Olaf Wolkenhauer

**Author notes:** Address of correspondence: S Bej, University of Rostock, Dept. of Systems Biology & Bionformatics, Universitätsplatz 1, 18051, Rostock, Germany. Address of correspondence: J Sarkar, CSIR-Indian Institute of Chemical Biology, 4 Raja SC Mullick Road, Kolkata 700032, India. Address of correspondence: O Wolkenhauer, University of Rostock, Dept. of Systems Biology & Bionformatics, Universitätsplatz 1, 18051, Rostock, Germany. first author(s) with equal contributions (Saptarshi Bej, Jit Sarkar).

## Abstract

**Background:** Studies on Type 2 Diabetes Mellitus (T2DM) have revealed heterogeneous sub-populations in terms of underlying pathologies. However, identification of subpopulations in epidemiological datasets remain unexplored. We here focus on the detection of T2DM clusters in epidemiological data, specifically analysing the National Family Health Survey-4 (NFHS-4) dataset containing a wide spectrum of features, including medical history, dietary and addiction habits, socio-economic and lifestyle patterns of 10,125 T2DM patients.

**Methods:** Epidemiological data provide challenges for analysis due to the diverse types of features in it. In this case, applying the state-of-the-art dimension reduction tool UMAP conventionally was found to be ineffective for the NFHS-4 dataset, which contains continuous, ordinal and nominal feature types. Continuous features, although smaller in numbers, had an overpowering effect on the distribution of clusters. We implemented a distributed clustering workflow combining different similarity measure settings of UMAP, for clustering continuous, ordinal and nominal features separately. We integrated the reduced dimensions from each feature-type-distributed clustering to obtain interpretable and unbiased clustering of the data.

**Findings:** From a methodological perspective, we show that for diverse data types, frequent in epidemiological datasets, feature-type-distributed clustering using UMAP is effective as opposed to the conventional use of the UMAP algorithm. Application of UMAP based clustering workflow for this type of dataset is novel in itself.

Our analysis reveals four significant clusters, with two of them comprising mainly of non-obese T2DM patients. These non-obese clusters has lower mean age and majorly comprises of rural residents. Surprisingly, one of the obese clusters had 90% of the T2DM patients practising non-vegetarian diet though they did not show an increased intake of plant-based protein-rich foods.

**Interpretation:** Our findings demonstrate the presence of a heterogeneity among T2DM patients with regard to socio-demography and dietary pattern. From our analysis, we conclude that, existence of significant non-obese T2DM subpopulations characterized by younger age group and economic disadvantage, raise the need of different screening criteria for T2DM among rural Indian residents.

**Funding:** This work was in part supported by funds from Bioinformatics Infrastructure (de.NBI) and Establishment of Systems Medicine Consortium in Germany e:Med, as well as the German Federal Ministry for Education and Research (BMBF) programs (FKZ 01ZX1709C). The work has also been funded and supported by the Indian Council of Medical research (ICMR) (No.3/1/3/JRF-2017/HRD-LS/56429/54).

## 1 Introduction

Type 2 Diabetes Mellitus (T2DM) is a multifactorial disease globally estimated to rise to 629 million cases by 2045 (See [IDF Diabetes Atlas](https://www.diabetesatlas.org/en/)) [1, 2]. Though conceived as a homogeneous disease for long, several recent studies have found T2DM to be a mix of heterogenous disease subtypes [3, 4, 5]. These studies have reported a varied pathophysiology underlying T2DM and thereby suggest the possibility of a personalised treatment for T2DM.

Besides obesity, other factors like age, sex, socio-economic status, place of residence (rural/urban), smoking habit, alcohol intake, food frequency etc. significantly associate with T2DM [6, 7, 8, 9, 10, 11, 12, 13]. Several of these factors are modifiable in nature and hence are important in the management of T2DM [1]. However, modification of lifestyle-related factors vary and thereby lead to a differential degree of glycemic control among T2DM patients [14]. Glycaemic control and response to anti-diabetics has also been shown to be different among T2DM sub-groups [15]. To explore whether any particular pattern of patient sub-populations exist within the entire T2DM population based on socio-demographic and lifestyle factors, we used an unsupervised clustering approach on the largest and most comprehensive epidemiological dataset in India, the National Family Health Survey-4 (NFHS-4) dataset. Clusters were subsequently characterised to identify unique socio-demographic and lifestyle patterns associated with these sub-populations.

Epidemiological datasets provide a comprehensive set of information regarding socio-demography, lifestyle, addiction and co-morbidities. Variables containing such information are called *features* in the language of Machine Learning. In the T2DM-NFHS-4 dataset, there are 36 such features, containing information on each diabetes patient. Moreover, in our dataset, the features can be categorised into three types:

1. *Continuous features:* These are the features which can assume any numeric value from a continuous range. For example, BMI of a patient is a continuous feature.
2. *Ordinal features:* These are the features which assume values from a discrete range, such that, there is a sense of order in the values assumed by the feature. For example, let us assume a feature ‘meat consumption by a patient’, assumes values ‘daily’, ‘weekly’ or ‘monthly’. Clearly the range of the feature ‘meat consumption by a patient’ is discrete, since it can assume any one of the three values. Also, there is a sense of order in the values, indicating that daily meat consumption is the highest and daily meat consumption is the lowest, if we want to quantify meat consumption.
3. *Nominal features:* These are the features which assume values from a discrete range, such that, there is no sense of order in the values assumed by the feature. For example, let us assume a feature ‘Religion of a patient’, assumes values ‘Hindus’, ‘Muslims’ or ‘Christians’. Clearly the range of the feature ‘meat consumption by a patient’ is discrete, since it can assume any one of the three values. But there is no sense of order in the possible values assumed by the features. Yet, this feature draws its importance from the fact that lifestyle patterns or diets vary largely among these religious groups.

Such diverse types of features in epidemiological data create challenges for the analysis. Conventional application of the state-of-the-art dimension reduction tool Uniform Manifold Approximation (UMAP) was found to be ineffective for the T2DM-NFHS-4 dataset,. Continuous features, although smaller in numbers, had a overpowering effect on the distribution of clusters. To address this problem, we implemented a distributed clustering workflow, combining different similarity measure settings of UMAP, for clustering continuous, ordinal and nominal features separately. We integrated the reduced dimensions from each feature-type-distributed clustering to obtain interpretable and unbiased clustering of the data.

The workflow realised for the present study (Figure 1) involves investigation of underlying socio-demographic patterns within patient sub-populations using unsupervised learning. Dimension reduction approaches are often used to reduce higher dimensional data to lower dimensions such that in the lower dimensional embedding of the data one can visualize underlying clusters within the data, that are not apparent in the higher dimensions [16]. Several such techniques have been developed over the last few decades. Until recently the dimension reduction technique t-Stochastic Neighbourhood Embedding (t-SNE) was a state-of the-art algorithm in this field providing numerous applications in various fields [17, 18, 19]. t-SNE projects high dimensional data to a lower dimension while maintaining the underlying local manifold structure in a sense that, in a lower dimension t-SNE can cluster points, that are close enough in the latent high dimensional manifold [17].

**Figure 1:**
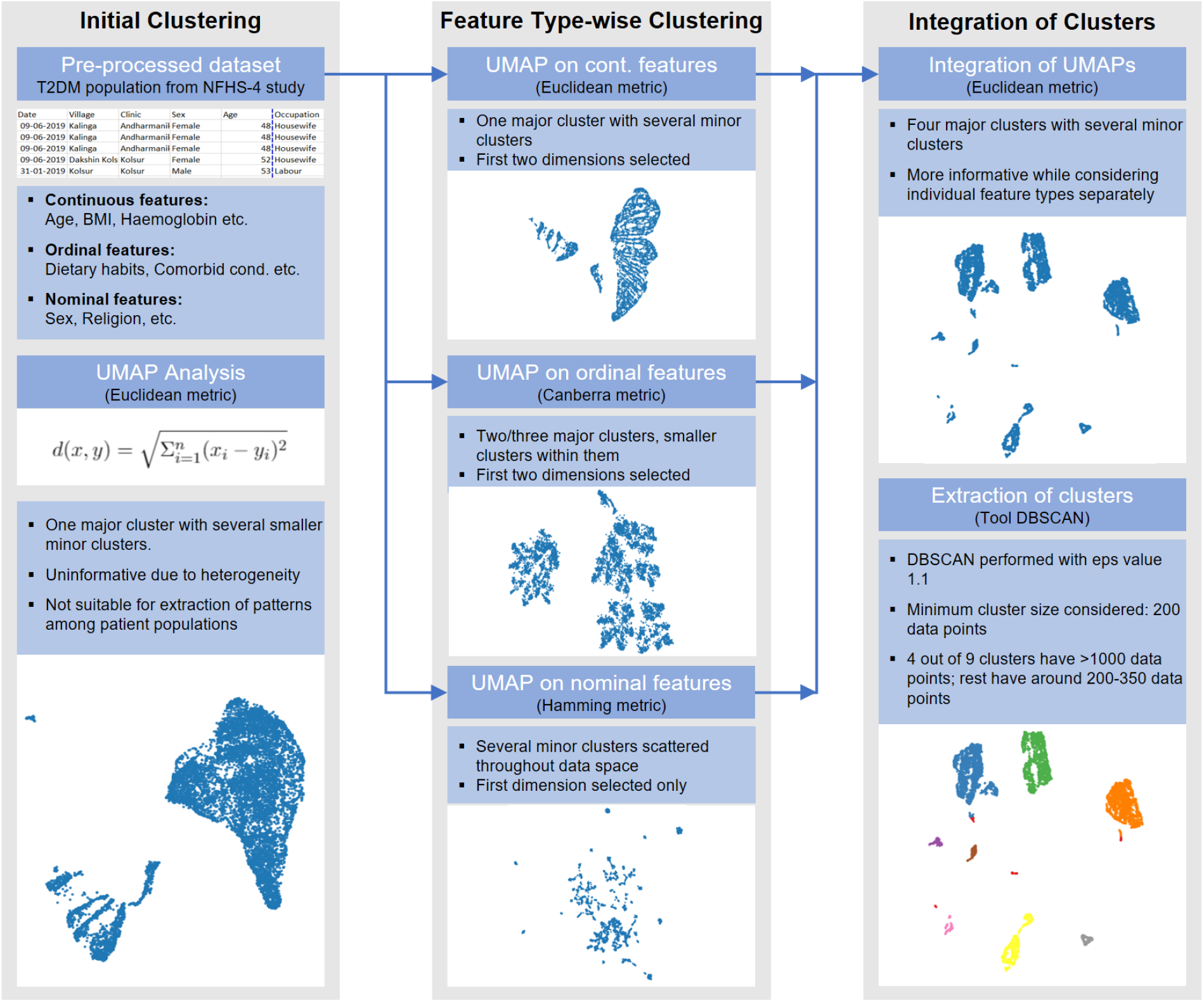
Workflow describing the analysis of the T2DM NFHS-4 Dataset.

With a rigorous mathematical foundation, considerably high speed and easy to use using scikitlearn API, UMAP has turned out to be one of the most popular choices among the data scientists [20, 21, 22]. As opposed to t-SNE, UMAP uses a graph based manifold approximation mechanism which contributes to preservation of the global as well as Social properties of the latent data manifold in a lower dimensional representation of the data. Given some low dimensional representation of the data, a similar process can be used to construct an equivalent topological representation. UMAP builds a graph considering customized neighbourhoods for every data points. This graph is a representation of the higher dimensional data manifold. The end result is a patchwork of low-dimensional representations of neighbourhoods that groups similar data points on a local scale while better preserving long-range topological connections to more distantly related data points [20, 22]. For the ability of UMAP to preserve the long-range topological connections along with the short-range topological connections and because of its high computational efficiency we choose UMAP for our unsupervised clustering approach. Moreover, UMAP allows an user to specify several similarity measures through the tuning of the `metric` parameter. This has been critical in our workflow, since our data contains continuous and categorical features and choosing suitable similarity measures for continuous and categorical features is crucial for a meaningful and informative clustering [23].

## 2 Methodology

### 2.1 Source and Description of the T2DM NFHS-4 Dataset

Data preparation and pre-processing are the key aspects of approaching a problem from a Machine Learning perspective. In this Section we provide the details on the pre-processing approach adopted to generate the T2DM-NFHS-4 dataset.

The NFHS-4 dataset was downloaded from [The Demographic & Health Surveys (DHS) Program website](https://www.dhsprogram.com/). NFHS-4 is the fourth version of national health survey conducted under the supervision of Ministry of Health and Family Welfare, Government of India with the International Institute for Population Sciences (IIPS), Mumbai serving as the main nodal agency for all the surveys. The sampling procedure followed in NFHS-4 was of stratified two-stage sampling covering all the 640 districts of India. The survey was successfully conducted with 601,509 households. In those interviewed households 112,122 men and 699,686 women could be successfully interviewed. Four survey questionnaires (Household Questionnaire, Woman’s Questionnaire, Man’s Questionnaire and Biomarker Questionnaire) were implemented in 17 local languages to collect information on basic demographic information, socio-economic parameters, family planning issues, nutritional status, health indicators, contact with community health workers etc. Uniqueness of the NFHS-4 study was that it collected data on Diabetes status and performed a Random Blood Glucose for individuals (15-54 years) using a finger-stick blood specimen. As a result, the biomarker measurements and tests besides anthropometric measurements like anaemia testing, blood pressure measurement, blood glucose testing and HIV testing were included in the survey.

### 2.2 Dataset Preparation

For dataset preparation and cleaning, the three questionnaires were merged-Woman’s Questionnaire, Man’s Questionnaire and Biomarker Questionnaire. The first two contained information about background characteristics (location, age, sex, religion, social group, literacy, wealth status etc), nutritional practices, addictions and co-morbidities while the bio-marker questionnaire contained information on height, weight, blood pressure and random blood glucose. A unique code was generated for all individuals in all the three questionnaires by appending the Country code and phase, Cluster number, Household number and Line number. The three datasets were joined by the unique code to prepare a single dataset of 810,971 individuals consisting of all men and women between 15-54 years of age. Pregnant women were next excluded to discard the possibility of Gestational Diabetes Mellitus. Individuals with missing diabetic and blood pressure status were also excluded. Variables known to be risk factors for DM (BMI, Age, Place of residence, Wealth Index, Smoking frequency, Alcohol intake frequency, Hypertension), socio-economic factors (Sex, Religion, Social group, Educational status), Dietary frequencies and haemoglobin level were selected for final analysis. BMI, age and haemoglobin level were taken as continuous variables and the rest as categorical variables. Outliers were removed separately for all the three continuous variables to obtain the final dataset with 610498 individuals (526678 females and 83820 males).

### 2.3 Dataset Preprocessing

We were interested in detecting significant T2DM sub-populations in the data and further sought to characterize these subpopulations based on the socio-demographic and co-morbid conditions. For this purpose, we extracted patients with known history of diabetes from the dataset: a total of 10,125 patients. We considered a diverse collection of socio-demographic and co-morbid conditions as ‘features’ in our dataset. Qualitatively our features can be divided into several categories:

1. *Co-morbid conditions:* This class of features considers the co-morbid diseases among T2DM patients. We considered whether a T2DM patient had medical conditions such as Asthma, Thyroid disorder, Heart disease, Cancer, Tuberculosis and Hypertension. Thus, there were six features in this category. These features are binary in nature denoting whether a T2DM patient suffered from a given comorbidity or not.
2. *Food habits:* This class of features considered the food habits of T2DM patients. The features considered here were how frequently the patient took the food items: Milk or Curd, Pulses or Beans, Dark leafy vegetables, Fruits, Eggs, Fish, Chicken, Fried food and Aerated drinks. Thus, there were nine features in this category. Features were categorical and ordinal in nature having four possible values: ‘Daily’, ‘Occasionally’, ‘Weekly’ and ‘Never’.
3. *Addiction history:* This class of features considered the addiction pattern of T2DM patients. There were two features in this class, both binary in nature encoding whether a patient is a Smoker or whether a patient takes Alcohol.
4. *Socio-demographic features:* These included features such as Sex, Age, Wealth index, Education level, Religion and Caste along with Body Mass Index (BMI) and Haemoglobin level of the patient. There were eight features in this category.
5. *Living conditions:* This class of features quantify the living conditions of the patients. The features in this class considered whether a patient lives in a household possessing refrigerator, bicycle, motorbike, four wheeler vehicle and livestock. Moreover, there were features denoting type of residence, household structure, frequency of household members smoking inside the house, type of cooking fuel used, source of drinking water and time to reach the nearest drinking water source. Thus, there were eleven features belonging to this category.

For our study, 36 features or factors are considered to investigate significant patient populations among the diabetes patients into consideration. Note that there are both continuous and categorical features among these thirty six features. Among the categorical features there are both ordinal features and nominal features. Ordinal features have a sense of order among them, such as the features from the ‘food habits’ category as described before. The nominal features are categorical features with no sense of order such as sex of a patient. Note that for our dataset the continuous features are: Age, BMI, Haemoglobin level and Time to get to drinking water source; whereas the nominal features are: Sex, Religion, Caste, Household structure, Type of place of residence, Type of cooking fuel and Source of drinking water. The rest of the features are ordinal features. The categorization of features into continuous, nominal and ordinal is of utmost importance in our clustering paradigm which we discuss in Section 2.4.1.

### 2.4 Identification of T2DM sub-populations using U-MAP and DBSCAN

From our detailed description of our dataset we pointed out that our dataset has a variety of features including continuous and categorical features. Further, there are both ordinal and nominal features among the categorical features in our dataset. A simple UMAP on the entire dataset is depicted in Figure 2(a), revealing two broad clusters. For this clustering UMAP parameters `n_neighbours` have been chosen to be 30, whereas the metric parameter has been chosen to be euclidean. However we have a number of important nominal and ordinal categorical features whose effect would not be apparent from such a clustering. Moreover, the euclidean distance does not always make sense on categorical features, especially if they are nominal in nature. For example, observe Figure 2(d), where we have used UMAP considering only the nominal features with metric parameter hamming (based on hamming distance). This reveals a completely different picture of the dataset, showing several small clusters. Our clustering paradigm is designed to optimise this effect and find a balance in the clustering where a particular type of feature does not have an overpowering effect on the clustering process.

**Figure 2:**
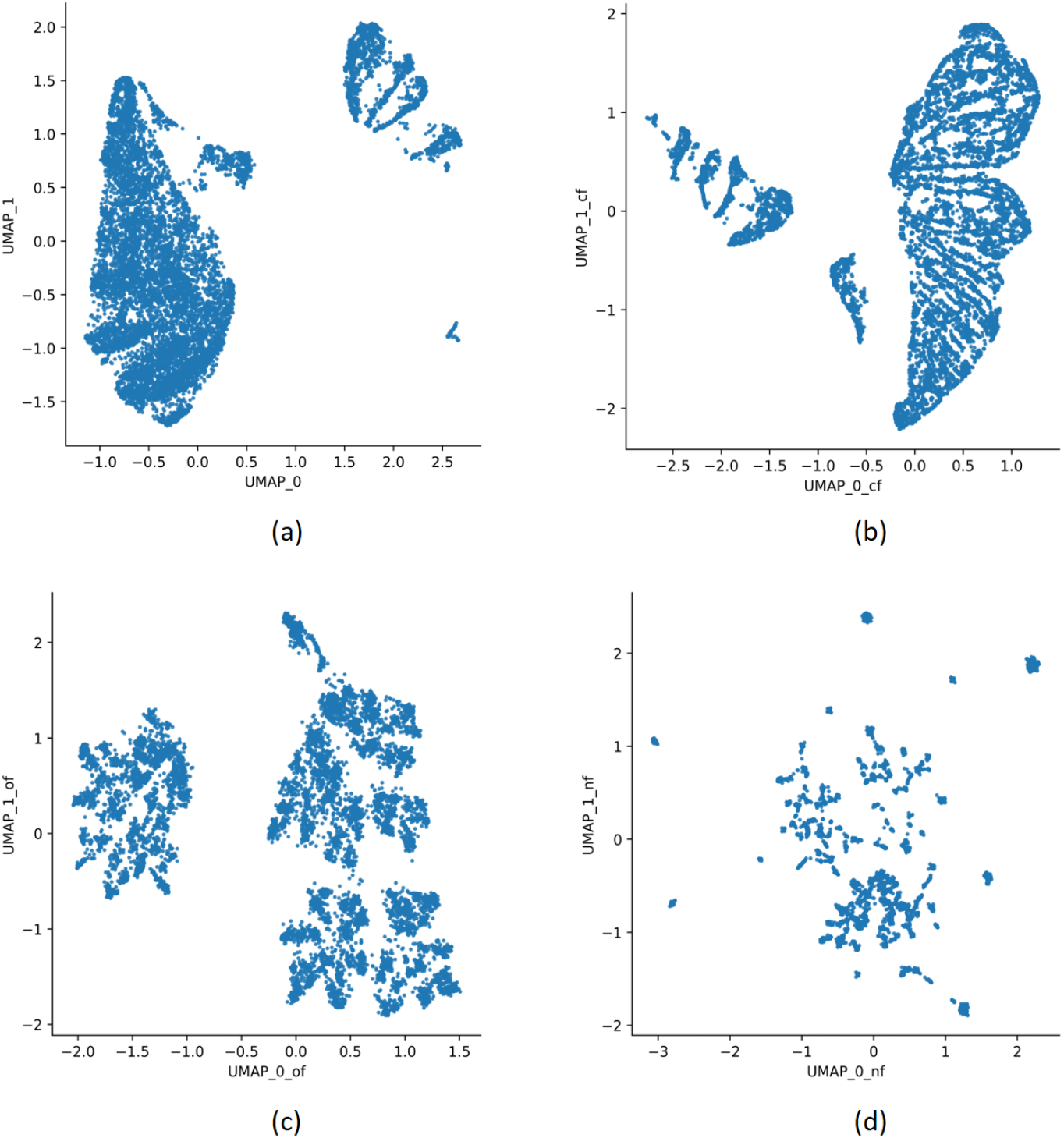
(a) Figure showing UMAP clusters for all the features with Euclidean metric (b) Figure showing UMAP clusters for continuous features with Euclidean metric (c) Figure showing UMAP clusters for ordinal features with Canberra metric (d) Figure showing UMAP clusters for nominal features with Hamming metric

### 2.4.1 Clustering paradigm using UMAP

Our clustering paradigm applies UMAP separately on continuous, nominal and ordinal features separately. For each of these feature categories we create a lower dimensional embedding of the dataset. Finally we integrate the lower dimensional embeddings to extract clusters from them using the DBSCAN algorithm, a clustering algorithm used for extracting clusters from data based on data density. One advantage of this algorithm is that one does not need to specify the number of clusters from beforehand. DBSCAN considers closely or densely located points, as clusters [24]. For UMAP, we use the same values for the parameters `n_neighbours`= 30 and `min_distance`= 0.1 for all the feature types.

- For the *continuous features* we use the `metric` measure to be *Euclidean*. The Euclidean distance between two vectors is given by:

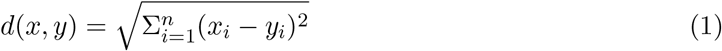
- For the *nominal features* we use the`metric` measure to be *Hamming*. Hamming distance is defined as:

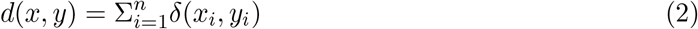

where *δ*(*x*_*i*_, *y*_*i*_) = 1 if *x*_*i*_ = *y*_*i*_ and *δ*(*x*_*i*_, *y*_*i*_) = 0 otherwise. Recall that, nominal features are also a type of categorical features which do not have a sense of order associated to them. For such features Hamming distance is widely used as a similarity measure between data points [23].
- For the *ordinal features* we use the `metric` measure to be *Canberra*. It is a weighted version of the Manhattan measure. The Canberra distance is given by:

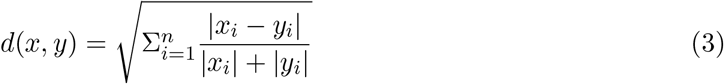

Ordinal features are also a type of categorical features. However, the Hamming metric can not capture the inherent ordered relationships and statistic information from categorical values [23]. We thus tried using UMAP for several metric measures and noticed that the Canberra distance measure retains a high variance in the lower dimensions. Thus we chose the Canberra distance measure as a similarity metric for ordinal features.

For the categorical and ordinal features we thus produce a two dimensional representation of each data point by taking into consideration the first two UMAP coordinates. For the nominal features we consider we produce a one dimensional representation, since the data points are too scattered in this case as shown in Figure 2(d) and thus can lead to too many clusters. Thus, we reduce every data point into a five dimension representation, two for each of the continuous and ordinal features and one for the nominal features. Finally, we look for clusters in the five dimensional representation using DBSCAN (`eps`= 1, `minpoints`= 200). After selecting the final clusters, we characterized them by summarizing all the 36 variables separately for each cluster. The continuous variables were summarized as their mean and the standard error of the mean. The categorical variables were summarized as their frequency distribution and the proportion of each value within each cluster.

#### 2.4.2 Extraction of T2DM sub-populations using DBSCAN

Using our clustering paradigm described before, we can detect seven subpopulations among the patients where 261 patients are considered as outliers. We show the distribution of clusters in Figure 3a. We further perform a UMAP on the five dimensional reduced representation of our data to visualize the clusters detected by DBSCAN. For this we label the data points using the DBSCAN clustering labels and colour code them in the UMAP representation of the five dimensional reduced data as shown in Figure 3b. This provides validation to the fact the clustering done by DBSCAN makes sense. Note that, from our clusters we can detect four significant patient subpopulations containing 2898, 2301, 2226 and 1315 data points.

**Figure 3:**
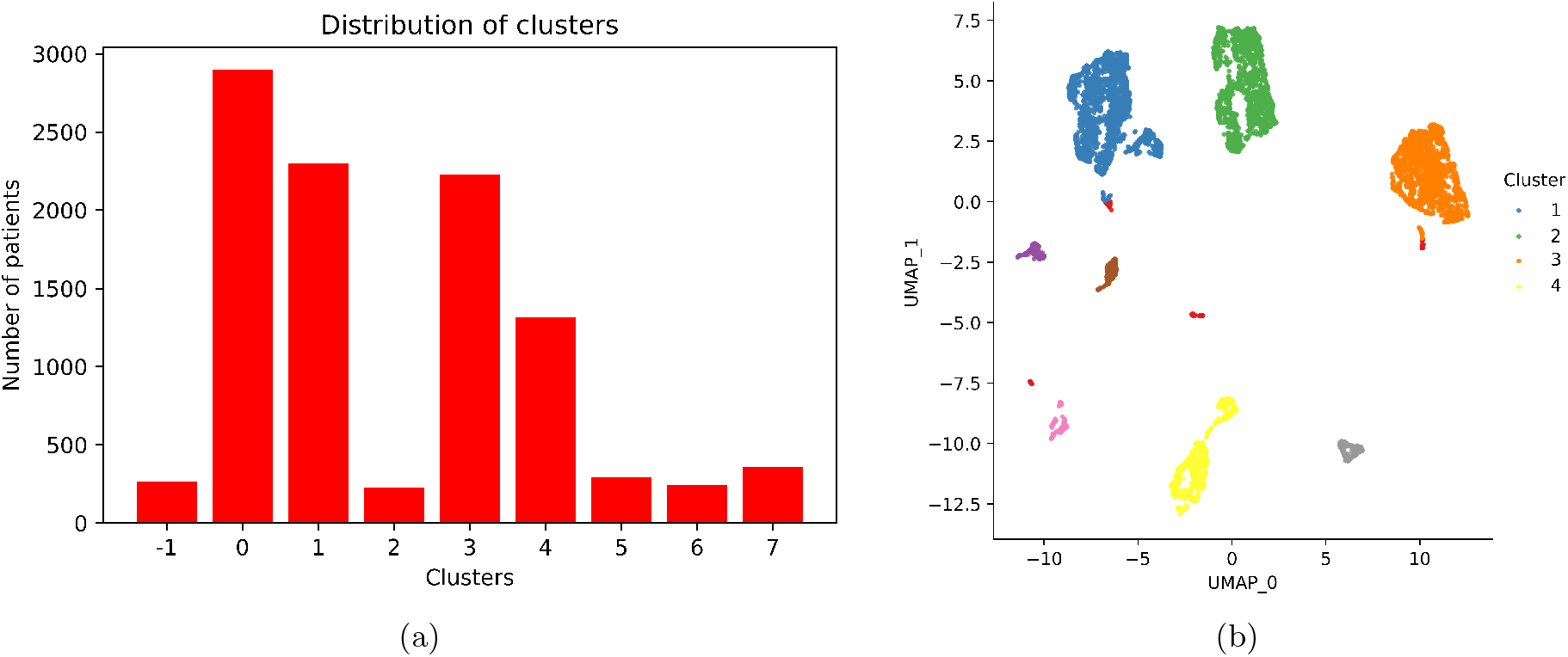
(a) Distribution of clusters detected by DBSCAN on the five dimensional reduced representation of the data (b) UMAP clusters for five dimensional reduced representation of the data annotated by the DBSCAN generated clusters

## 3 Results

### 3.1 Characterization of clusters

#### Age and BMI both were found to be lower in Cluster 2 and Cluster 4

Age and obesity are the most important risk factors for T2DM. However, we found a heterogeneity in both these variables across all the clusters. Interestingly, the mean Age and BMI both were lower in Cluster 2

(Age: 38.3 ± 0.19 years, BMI: 23.9 ± 0.1) and Cluster 4 (Age: 37.9 ± 0.26 years, BMI: 23.6 ± 0.13) compared to Cluster 1 (Age: 41.3 ± 0.14 years, BMI: 26.7 ± 0.09) and Cluster 3 (Age: 39.9 ± 0.18 years, BMI: 26 ± 0.11). However distribution of males and females has been found to be similar across all the clusters.

#### Higher proportion of rural residents and lower proportion of richest wealth quintile in Cluster 2 and 4

Proportion of rural residents was found to be high in Cluster 2 (69.4% were Rural residents) and Cluster 4 (72.02% were Rural residents) compared to the other clusters (31.3% in

Cluster 1 and 49.19% in Cluster 3). Surprisingly, only 4.3% people in Cluster 2 and 8.37% in Cluster 4 belonged to the richest quintile of the Wealth Index category whereas 64.04% in Cluster 1 and 54.9% in Cluster 3 belonged to the same.

#### Frequency of co-morbid conditions were similar across all the clusters

Co-morbid conditions included history of asthma, thyroid disease, heart disease, cancer, history of tuberculosis, haemoglobin level and hypertension. Though the distribution of disease conditions show minor variation across the clusters (Table 1), the trend is almost similar in all the clusters.

**Table 1:** Detailed cluster-specific analysis for all numerical and categorical variables.

| Identified clusters |  | Cluster 1 | Cluster 2 | Cluster 3 | Cluster 4 |
| --- | --- | --- | --- | --- | --- |
| Cluster Size (N) |  | 2898 | 2301 | 2226 | 1315 |
| Cont. Variables (Mean ± SE) | | | | | |
| Age (yrs) | | 41.3 ± 0.14 | 38.3 ± 0.19 | 39.9 ± 0.18 | 37.9 ± 0.26 |
| Body Mass Index (kg/m<sup>2</sup>) | | 26.7 ± 0.09 | 23.9 ± 0.1 | 26 ± 0.11 | 23.6 ± 0.13 |
| Haemoglobin (gm/dl) | | 12.5 ± 0.04 | 12.3 ± 0.04 | 12.1 ± 0.04 | 12.3 ± 0.06 |
| Time to Water Source (min) | | 0.1 ± 0.01 | 0.02 ± 0.01 | 0.09 ± 0.01 | 18.6 ± 0.39 |
| Cat.Variables | Value for cat. variables |  |  |  |  |
| Sex | Male | 558 (19.25) | 457 (19.86) | 270 (12.13) | 323 (24.56) |
|  | Female | 2340 (80.75) | 1844 (80.14) | 1956 (87.87) | 992 (75.44) |
| History of Asthma | No | 2737 (94.44) | 2064 (89.7) | 1999 (89.8) | 1121 (85.25) |
|  | Yes | 161 (5.56) | 237 (10.3) | 227 (10.2) | 194 (14.75) |
| History of Thyroid Disorder | No | 2636 (90.96) | 2135 (92.79) | 1992 (89.49) | 1196 (90.95) |
|  | Yes | 262 (9.04) | 166 (7.21) | 234 (10.51) | 119 (9.05) |
| History of Heart Disease | No | 2729 (94.17) | 2107 (91.57) | 1996 (89.67) | 1174 (89.28) |
|  | Yes | 169 (5.83) | 194 (8.43) | 230 (10.33) | 141 (10.72) |
| History of Cancer | No | 2876 (99.24) | 2272 (98.74) | 2161 (97.08) | 1246 (94.75) |
|  | Yes | 22 (0.76) | 29 (1.26) | 65 (2.92) | 69 (5.25) |
| Ever suffered from TB | No | 2890 (99.72) | 2287 (99.39) | 2218 (99.64) | 1305 (99.24) |
|  | Yes | 8 (0.28) | 14 (0.61) | 8 (0.36) | 10 (0.76) |
| Milk/Curd intake freq | Never | 201 (6.94) | 183 (7.95) | 110 (4.94) | 123 (9.35) |
|  | Weekly | 461 (15.91) | 551 (23.95) | 293 (13.16) | 405 (30.8) |
|  | Occasionally | 611 (21.08) | 669 (29.07) | 447 (20.08) | 291 (22.13) |
|  | Daily | 1625 (56.07) | 898 (39.03) | 1376 (61.81) | 496 (37.72) |
| Pulses/Beans intake freq | Never | 13 (0.45) | 17 (0.74) | 18 (0.81) | 9 (0.68) |
|  | Weekly | 255 (8.8) | 248 (10.78) | 152 (6.83) | 198 (15.06) |
|  | Occasionally | 1263 (43.58) | 937 (40.72) | 936 (42.05) | 574 (43.65) |
|  | Daily | 1367 (47.17) | 1099 (47.76) | 1120 (50.31) | 534 (40.61) |
| Green vegetables intake freq | Never | 7 (0.24) | 12 (0.52) | 10 (0.45) | 9 (0.68) |
|  | Weekly | 324 (11.18) | 259 (11.26) | 279 (12.53) | 142 (10.8) |
|  | Occasionally | 1000 (34.51) | 796 (34.59) | 792 (35.58) | 483 (36.73) |
|  | Daily | 1567 (54.07) | 1234 (53.63) | 1145 (51.44) | 681 (51.79) |
| Fruit intake freq | Never | 50 (1.73) | 65 (2.82) | 74 (3.32) | 41 (3.12) |
|  | Weekly | 897 (30.95) | 1148 (49.89) | 872 (39.17) | 750 (57.03) |
|  | Occasionally | 1203 (41.51) | 818 (35.55) | 810 (36.39) | 386 (29.35) |
|  | Daily | 748 (25.81) | 270 (11.73) | 470 (21.11) | 138 (10.49) |
| Egg intake freq | Never | 97 (3.35) | 85 (3.69) | 1983 (89.08) | 41 (3.12) |
|  | Weekly | 1005 (34.68) | 963 (41.85) | 153 (6.87) | 520 (39.54) |
|  | Occasionally | 1537 (53.04) | 1100 (47.81) | 80 (3.59) | 678 (51.56) |
|  | Daily | 259 (8.94) | 153 (6.65) | 10 (0.45) | 76 (5.78) |
| Fish intake freq | Never | 222 (7.66) | 106 (4.61) | 2162 (97.12) | 83 (6.31) |
|  | Weekly | 994 (34.3) | 1006 (43.72) | 35 (1.57) | 593 (45.1) |
|  | Occasionally | 1210 (41.75) | 987 (42.89) | 20 (0.9) | 563 (42.81) |
|  | Daily | 472 (16.29) | 202 (8.78) | 9 (0.4) | 76 (5.78) |
| Chicken/Meat intake freq | Never | 53 (1.83) | 58 (2.52) | 2175 (97.71) | 33 (2.51) |
|  | Weekly | 1274 (43.96) | 1150 (49.98) | 32 (1.44) | 640 (48.67) |
|  | Occasionally | 1475 (50.9) | 1032 (44.85) | 18 (0.81) | 612 (46.54) |
|  | Daily | 96 (3.31) | 61 (2.65) | 1 (0.04) | 30 (2.28) |
| Fried food intake freq | Never | 179 (6.18) | 161 (7) | 276 (12.4) | 95 (7.22) |
|  | Weekly | 1275 (44) | 988 (42.94) | 1114 (50.04) | 631 (47.98) |
|  | Occasionally | 1071 (36.96) | 849 (36.9) | 715 (32.12) | 408 (31.03) |
|  | Daily | 373 (12.87) | 303 (13.17) | 121 (5.44) | 181 (13.76) |
| Aerated drink intake freq | Never | 512 (17.67) | 475 (20.64) | 409 (18.37) | 262 (19.92) |
|  | Weekly | 1579 (54.49) | 1258 (54.67) | 1200 (53.91) | 744 (56.58) |
|  | Occasionally | 597 (20.6) | 449 (19.51) | 497 (22.33) | 236 (17.95) |
|  | Daily | 210 (7.25) | 119 (5.17) | 120 (5.39) | 73 (5.55) |
| Alcoholic | No | 2627 (90.65) | 2027 (88.09) | 2171 (97.53) | 1127 (85.7) |
|  | Yes | 271 (9.35) | 274 (11.91) | 55 (2.47) | 188 (14.3) |
| Smoker | No | 2770 (95.58) | 2192 (95.26) | 2197 (98.7) | 1234 (93.84) |
|  | Yes | 128 (4.42) | 109 (4.74) | 29 (1.3) | 81 (6.16) |
| Indoor Smoking freq | Never | 1849 (63.8) | 1138 (49.46) | 1429 (64.2) | 690 (52.47) |
|  | Weekly | 222 (7.66) | 264 (11.47) | 176 (7.91) | 129 (9.81) |
|  | Less than monthly | 72 (2.48) | 72 (3.13) | 71 (3.19) | 33 (2.51) |
|  | Monthly | 78 (2.69) | 72 (3.13) | 68 (3.05) | 36 (2.74) |
|  | Daily | 677 (23.36) | 755 (32.81) | 482 (21.65) | 427 (32.47) |
| Residence | Urban | 1991 (68.7) | 704 (30.6) | 1131 (50.81) | 368 (27.98) |
|  | Rural | 907 (31.3) | 1597 (69.4) | 1095 (49.19) | 947 (72.02) |
| Wealth Index | Poorest | 1 (0.03) | 287 (12.47) | 82 (3.68) | 301 (22.89) |
|  | Poorer | 8 (0.28) | 519 (22.56) | 154 (6.92) | 285 (21.67) |
|  | Middle | 151 (5.21) | 698 (30.33) | 245 (11.01) | 339 (25.78) |
|  | Richer | 882 (30.43) | 698 (30.33) | 523 (23.5) | 280 (21.29) |
|  | Richest | 1856 (64.04) | 99 (4.3) | 1222 (54.9) | 110 (8.37) |
| Highest Education level | No education | 388 (13.39) | 758 (32.94) | 416 (18.69) | 472 (35.89) |
|  | Primary level | 347 (11.97) | 373 (16.21) | 303 (13.61) | 240 (18.25) |
|  | Secondary level | 1641 (56.63) | 1006 (43.72) | 1106 (49.69) | 530 (40.3) |
|  | Higher level | 522 (18.01) | 164 (7.13) | 401 (18.01) | 73 (5.55) |
| Religion | Hindu | 1822 (62.87) | 1544 (67.1) | 1947 (87.47) | 975 (74.14) |
|  | Muslim | 627 (21.64) | 472 (20.51) | 46 (2.07) | 210 (15.97) |
|  | Christian | 313 (10.8) | 210 (9.13) | 13 (0.58) | 97 (7.38) |
|  | Others | 136 (4.69) | 75 (3.26) | 220 (9.88) | 33 (2.51) |
| Caste/Tribe | OBC | 1331 (45.93) | 871 (37.85) | 805 (36.16) | 472 (35.89) |
|  | SC | 384 (13.25) | 517 (22.47) | 328 (14.73) | 343 (26.08) |
|  | ST | 303 (10.46) | 385 (16.73) | 86 (3.86) | 258 (19.62) |
|  | General | 880 (30.37) | 528 (22.95) | 1007 (45.24) | 242 (18.4) |
| Blood Pressure | No | 1594 (55) | 1443 (62.71) | 1281 (57.55) | 849 (64.56) |
|  | Yes | 1304 (45) | 858 (37.29) | 945 (42.45) | 466 (35.44) |
| Possess Refrigerator | No | 131 (4.52) | 2296 (99.78) | 762 (34.23) | 989 (75.21) |
|  | Yes | 2767 (95.48) | 5 (0.22) | 1464 (65.77) | 326 (24.79) |
| Possess Bicycle | No | 1503 (51.86) | 1055 (45.85) | 1013 (45.51) | 617 (46.92) |
|  | Yes | 1395 (48.14) | 1246 (54.15) | 1213 (54.49) | 698 (53.08) |
| Possess Motorbike | No | 825 (28.47) | 1590 (69.1) | 734 (32.97) | 884 (67.22) |
|  | Yes | 2073 (71.53) | 711 (30.9) | 1492 (67.03) | 431 (32.78) |
| Possess Car/Truck | No | 2217 (76.5) | 2226 (96.74) | 1840 (82.66) | 1273 (96.81) |
|  | Yes | 681 (23.5) | 75 (3.26) | 386 (17.34) | 42 (3.19) |
| Cooking Fuel used | Other | 1 (0.03) | 4 (0.17) | 0 (0) | 1 (0.08) |
|  | Plant based | 354 (12.22) | 1018 (44.24) | 437 (19.63) | 723 (54.98) |
|  | Livestock based | 47 (1.62) | 297 (12.91) | 211 (9.48) | 104 (7.91) |
|  | Gas/Oil | 2460 (84.89) | 965 (41.94) | 1562 (70.17) | 476 (36.2) |
|  | Electricity | 36 (1.24) | 17 (0.74) | 16 (0.72) | 11 (0.84) |
| Household Structure | Non-nuclear | 1310 (45.2) | 1016 (44.15) | 1120 (50.31) | 564 (42.89) |
|  | Nuclear | 1588 (54.8) | 1285 (55.85) | 1106 (49.69) | 751 (57.11) |
| Possess Livestock | No | 2226 (76.81) | 1155 (50.2) | 1474 (66.22) | 646 (49.13) |
|  | Yes | 672 (23.19) | 1146 (49.8) | 752 (33.78) | 669 (50.87) |
| Drinking Water Source | Unprotrected sources | 76 (2.62) | 146 (6.35) | 44 (1.98) | 204 (15.51) |
|  | Protected sources | 739 (25.5) | 998 (43.37) | 686 (30.82) | 522 (39.7) |
|  | Community service | 1991 (68.7) | 1112 (48.33) | 1448 (65.05) | 508 (38.63) |
|  | Bottled water | 86 (2.97) | 43 (1.87) | 46 (2.07) | 77 (5.86) |
|  | Other | 6 (0.21) | 2 (0.09) | 2 (0.09) | 4 (0.3) |

#### Lifestyle patterns show evidences of a lower quality of life for patient sub-populations in Cluster 2 and 4

Our analysis reveal several other factors that support the fact that T2DM sub-populations from Cluster 2 and Cluster 4 have a considerably lower quality of life.

1. We observe that only 0.22% and 24.79% of patients belonging to Cluster 2 and Cluster 4 respectively possess a refrigerator compared to 95.48% and 65.77% of patients belonging to Cluster 1 and Cluster 3 respectively.
2. Only 30.9% and 32.78% of patients belonging to Cluster 2 and Cluster 4 respectively possess a motorbike compared to 71.53% and 67.03% of patients belonging to Cluster 1 and Cluster 3 respectively.
3. Only 3.26% and 3.19% of patients belonging to Cluster 2 and Cluster 4 respectively possess a car/truck compared to 23.5% and 17.34% of patients belonging to Cluster 1 and Cluster 3 respectively.
4. 44.24% and 54.98% of patients belonging to Cluster 2 and Cluster 4 respectively, use plant based cooking fuel, which is relatively cheap, compared to 12.22% and 19.63% of patients belonging to Cluster 1 and Cluster 3 respectively. Moreover, only 41.94% and 36.2% of patients belonging to Cluster 2 and Cluster 4 respectively use Gas/Oil based cooking fuel, which is relatively expensive, compared to 84.89% and 70.17% of patients belonging to Cluster 1 and Cluster 3 respectively.
5. 6.35 % and 15.51% of patients belonging to Cluster 2 and Cluster 4 respectively, drink water from unprotected sources, compared to 2.62% and 1.98% of patients belonging to Cluster 1 and Cluster 3 respectively.

#### Intake of non-vegetarian foods is invariably low in Cluster 3

Around 90% of the population in Cluster 3 had no intake of Egg (89.08%), fish (97.12%), chicken or meat (97.71%) whereas only less than 10% of the population in all the other 3 clusters had no intake of these non-vegetarian foods (Table 1). Though the Cluster 3 population had the highest daily intake of milk/curd (61.81%) and pulses/beans (50.31%) compared to the other clusters, other clusters also had almost similar proportion of people taking milk/curd and pulses/beans daily. Intake of other foods like dark leafy vegetables, fruits, fried foods and aerated drinks showed similar distribution across all the clusters.

## 4 Discussion

### 4.1 Rationale of the workflow in clustering epidemiological data

The clustering workflow used arises from some important observations that we will discuss here. To begin with we have a population of 10,125 T2DM patients with a diverse ensemble of features accounting for information on medical history, dietary and addiction habits, socio-economic and lifestyle patterns. Moreover, the features in the considered dataset are also diverse in terms of data types. We have a total of 36 features, out of which 4 are continuous features, 7 nominal features and 25 ordinal features, all of equal importance by assumption.

The aim is to find significant sub-populations in our data such that the identified sub-populations are interpretable in terms of the considered features. Note here that, by significant subpopulations we mean a subpopulation consisting of at least 10 percent of the total population. If there exists such sub-populations and we can explain the subpopulations in terms of the considered features, we can argue that these patterns exist in significant number of patients.

We have already argued in favour of using UMAP for our unsupervised approach to find clusters in the data. However, we observed that applying UMAP algorithm conventionally using the euclidean similarity metric on our entire dataset with 36 features turns out to be ineffective. The reason is, in this case the continuous features have an overpowering effect over the other feature types in determining the distribution of clusters. This can be observed from Figure 2(a) and 2(b). Note that Figure 2(a) shows UMAP clustering with all 36 features and 2(b) shows UMAP clustering with only four continuous features. Note that, there is a similarity in the clustering distribution of these figures, each containing one major cluster and seven small minor clusters. We observed that this is because of the fact that UMAP, when applied on all 36 features of the dataset using euclidean similarity measure is largely biased towards finding similarity among data points only in terms of the continuous features. Given that we have only four continuous features out of 36, this poses a problem as the diverse information present in the dataset in the form of the ordinal and nominal features are largely ignored.

To solve this problem, the clustering of continuous, ordinal and nominal features were treated separately by using different similarity matrices for them, giving rise to our clustering paradigm. We argued on our choice of similarity measures in Section 2.4.1. This generates for each feature type a data representation of lower dimension shown in Figure 2(b-d). We finally integrated these lower dimension data representations by taking two dimensional representations for continuous and ordinal features and an one dimensional representation (the one consisting of the most variance) for nominal features. The reason behind considering one dimensional representation for nominal features, is that using Hamming metrics for such data results in retaining a lot of variance in the data resulting in multiple clusters as we observe in Figure 2(d). Considering a two dimensional representation for this data while integrating these lower dimension data representations carry forward this variance and result in multiple small clusters in the final clustering distribution, which contradicts our aim of finding significantly large sub-populations (of at least 10 percent of the total population).

Finally, the integration is done by applying UMAP on the five dimensional reduced representation of the dataset using euclidean similarity measure (shown in Figure 3b). Note here that, in our final clusters we can observe patterns in all of continuous, ordinal and nominal data types. For example, in Cluster 4 the continuous feature ‘Time to Water source (min)’ shows very high values compared to other clusters. In Cluster 1 and 3, the nominal feature ‘Cooking fuel used’ shows a higher percentage for Gas/Oil users while in Cluster 2 and 4 the same feature shows a higher percentage for plant-based fuel users. In Cluster 3, the ordinal feature ‘Fish intake frequency’ shows a 97 percent of people to be never consuming fish. Thus, we infer that our clustering paradigm enables us to find significant sub-populations while keeping the clustering distribution unbiased, that is no feature type continuous, ordinal and nominal has an overpowering effect on the other.

### 4.2 Significance of T2DM clusters

T2DM was identified as a homogeneous disease with Insulin Resistance followed by *β*-cell dysfunction being the underlying pathology. However recent studies have explored and found T2DM to be a heterogeneous entity with the relative contribution of Insulin Resistance and *β*-cell dysfunction to differ across T2DM clusters [3]. These studies were performed on clinical and biochemical data with variables having uniform data types. On the other hand, our clustering approach takes into account the diverse data types obtained from an epidemiological dataset and discovers clusters among the T2DM population. Interestingly, two of the four clusters obtained in our study belonged to the non-obese T2DM phenotype where the mean BMI was below 25. These two non-obese clusters also had lower mean age compared to the other clusters. Both these non-obese clusters had larger proportion of rural residents and lower proportion of people belonging to the highest wealth quintile concluding to the fact that a large majority of T2DM people from rural India have lower BMI and are younger in age. The T2DM patient subpopulation belonging to these clusters have a relatively lower quality of life judging by analysis the lifestyle pattern based features. The non-obese phenotype of T2DM has been increasingly reported over the last two decades raising concern about the uniqueness of its underlying pathophysiology with a greater contribution of *β*-cell dysfunction compared to Insulin Resistance [25, 26, 27, 28]. This non-obese T2DM phenotype has been found among Asians and studies depicting and investigating its similarities and differences has been in place. Studies have concluded T2DM to occur among the Asians at a lower BMI cut-off and also at a younger age [29, 30]. This finding of two non-obese clusters with lower mean age provides confirmation to this.

Though non-obese T2DM is being considered as a unique phenotype, epidemiological studies for identifying high-risk population groups still remain undone. This is especially important for many Asian countries where over half of the T2DM population is of non-obese phenotype [25]. This analysis, reporting an increased presence of Rural residents in both the non-obese T2DM clusters, calls for a modification in BMI and Age cut-off for T2DM screening among rural residents. However identification of risk factors for T2DM specific to the rural population needs to be done. Representation of people from the highest wealth quintile was much lower in both the non-obese T2DM clusters. T2DM is a multi-factorial disease requiring strict compliance to lifestyle modification, proper diet and anti-diabetic therapy. Non-obese T2DM clusters with reduced representation from the highest wealth quintile suggests the possibility of an unequal access to care for non-obese T2DM people thereby generating the need of a more equitable healthcare policy in terms of prevention and therapy.

On the other hand, both the obese T2DM clusters had higher age and more urban residents. The proportion of people from the highest wealth quintile was higher in both the obese clusters. Interestingly one of the obese clusters (Cluster 3) had invariably low intake of non-vegetarian foods (egg, fish, chicken and meat) pointing out to the fact this T2DM cluster comprised of non-vegetarian people mainly. Dietary requirements in diagnosed T2DM patients involves reduced amount of carbohydrates and fats with increased amount of protein-rich foods [31]. Animal products, being rich sources of dietary protein, need to be included in the diet. One of the obese T2DM clusters with a strict non-vegetarian dietary pattern suggests the need to design a proper dietary guidelines for this group.

## 5 Conclusion

From a data science perspective, this analysis addresses the issue of diverse data types. We have shown that for such data conventional application of dimension reduction approaches might not be fruitful. We develop a workflow that contributes to finding meaningful and interpretable clusters such that the distribution of clusters is not biased by the data types.

Existence of a significant non-obese T2DM patient sub-population belonging to younger age group and having larger proportions of rural residents raises with a lower quality of life, indicate the need of a different screening criteria for T2DM among rural Indian residents. The obese T2DM cluster with around 90% of people sticking to the non-vegetarian diet calls for the need of dietary guidelines for T2DM patients having a non-vegetarian dietary pattern.

## Data Availability

We support the idea of transparency and reproducibility of research. Therefore, all data relevant to this work are made publicly available on a GitHub repository.

https://github.com/Saptarshi-Bej/Type-2-Diabetes-Mellitus-T2DM-/blob/master/Preprocessed_DM_xx.zip

## Data availability

We support the idea of transparency and reproducibility of research. Therefore, all data relevant to this work are made publicly available on the GitHub repository https://github.com/Saptarshi-Bej/Type-2-Diabetes-Mellitus-T2DM-/blob/master/Preprocessed_DM_xx.zip. More-over, the python code (in form of a jupyer notebook) for the implementation of our workflow is also provided publicly in https://github.com/Saptarshi-Bej/Type-2-Diabetes-Mellitus-T2DM-/blob/master/Clustering_paradigm_disc_cont.ipynb.

## Author Contributions

Saptarshi Bej and Jit Sarkar are the first authors and contributed equally to this work. Saptarshi Bej, Jit Sarkar, Pabitra Mitra, Partha Chakrabarti and Olaf Wolkenhauer contributed to the study concept and design. Saptarshi Bej, Jit Sarkar and Saikat Biswas did the data analysis. Saptarshi Bej, Jit Sarkar and Olaf Wolkenhauer wrote the manuscript and are the guarantors of this work having full access to all the data in the study and takes responsibility for the integrity of the data and the accuracy of the data analysis. All authors approved the final version of the article, including the authorship list.

## Acknowledgements

This work was in part supported by funds from Bioinformatics Infrastructure (de.NBI) and Establishment of Systems Medicine Consortium in Germany e:Med, as well as the German Federal Ministry for Education and Research (BMBF) programs (FKZ 01ZX1709C). JS received a research fellowship from Indian Council of Medical research (ICMR) (No.3/1/3/JRF-2017/HRD-LS/56429/54).

